# Generative Artificial Intelligence in Medical Education and Participatory Research for Social Action: A Human and AI Comparative Analysis

**DOI:** 10.64898/2026.05.14.26351842

**Authors:** Susana Juniu, Delivette Castor, Harry Reyes Nieva, Rita Charon, Silvia Amesty

## Abstract

Participatory qualitative methods such as Photovoice are increasingly used to link research with social action. Recent advances in artificial intelligence (AI) may enhance data analysis, inference, and action planning within such participatory approaches. This study explored medical students’ perceptions of social justice using conventional Photovoice analysis and assessed the potential contribution of generative AI (genAI). Nine students joined a six-week seminar, “Exploring the Concept of Social Justice Using Photovoice.” An initial two-hour session covered ethics, the Photovoice framework, and photography techniques. Participants then captured images reflecting their views on social justice, wrote narratives, and engaged in guided group discussions. Human researchers and students conducted a three-stage Photovoice analysis: 1) selecting photographs, 2) contextualizing them with participant narratives, and 3) inductively coding themes. To explore how AI might support data analysis, the research team analyzed the same data with five generative tools, including Sonix, ChatGPT, and Copilot. AI-generated themes and visual representations were compared with human-derived results for congruence, depth, and suggested action steps. Conventional analysis identified five major themes: (1) Social Justice and Inequality, (2) Contradictions and the Costs of Justice, (3) Community and Collective Action, (4) Environment and Environmental Justice, and (5) Perception, Subjectivity, and Perspective. AI-assisted analysis yielded six unified themes that closely aligned with human findings. Traditional Photovoice images conveyed authentic, lived experiences and strong emotional meaning, providing a powerful foundation for advocacy. AI-generated images and thematic summaries offered efficiency, creativity, and reduced researcher bias, improving generalizability. However, they lacked the emotional depth and contextual nuance present in participant-created visuals.

## 1 Introduction

Human insight and generative artificial intelligence (genAI) offer potentially distinct and complementary capabilities for qualitative analyses, including expanded interpretive capacity for contextual depth, meaning-making, and social action. This study sought to explore the use of genAI to enhance Photovoice as an educational tool for social justice among medical students.

### 1.1 Photovoice

Photovoice is a Community-Based Participatory Research (CBPR) methodology (**Figure 1**) that empowers participants to express themselves through photography and images, creating new opportunities for reflection on their lived experiences, promoting dialogue, and representing relevant community issues in a creative, critical, and personal manner, with the power to advocate for change (Wang, 1999). Photovoice is grounded in critical theory, offering a subjective perspective on reality through photographs representing participants’ self-reflections of their lived experiences (Freire, 1970). These visual testimonies and interpretations foster critical dialogue, reflective analysis, and pathways for social change (Riddick and Russell, 2015). Photovoice seeks to catalyze change for the betterment of the community by strengthening participants’ ability to think both logically and creatively (Budig et al., 2018). Photovoice aims to produce research and knowledge in collaboration with communities, ensuring that the knowledge produced is relevant to the community’s needs. Photovoice is a participatory methodology that combines critical pedagogy, feminist theory, and documentary photography to empower individuals, particularly those from marginalized communities, to examine, express, and act upon the social realities they face (Wang et al., 1996; Wang and Burris, 1997). Rooted in Freire’s concept of conscientization, a process that develops critical awareness through reflection and action, it seeks to shift power in knowledge production and advocacy.

**Figure 1.**
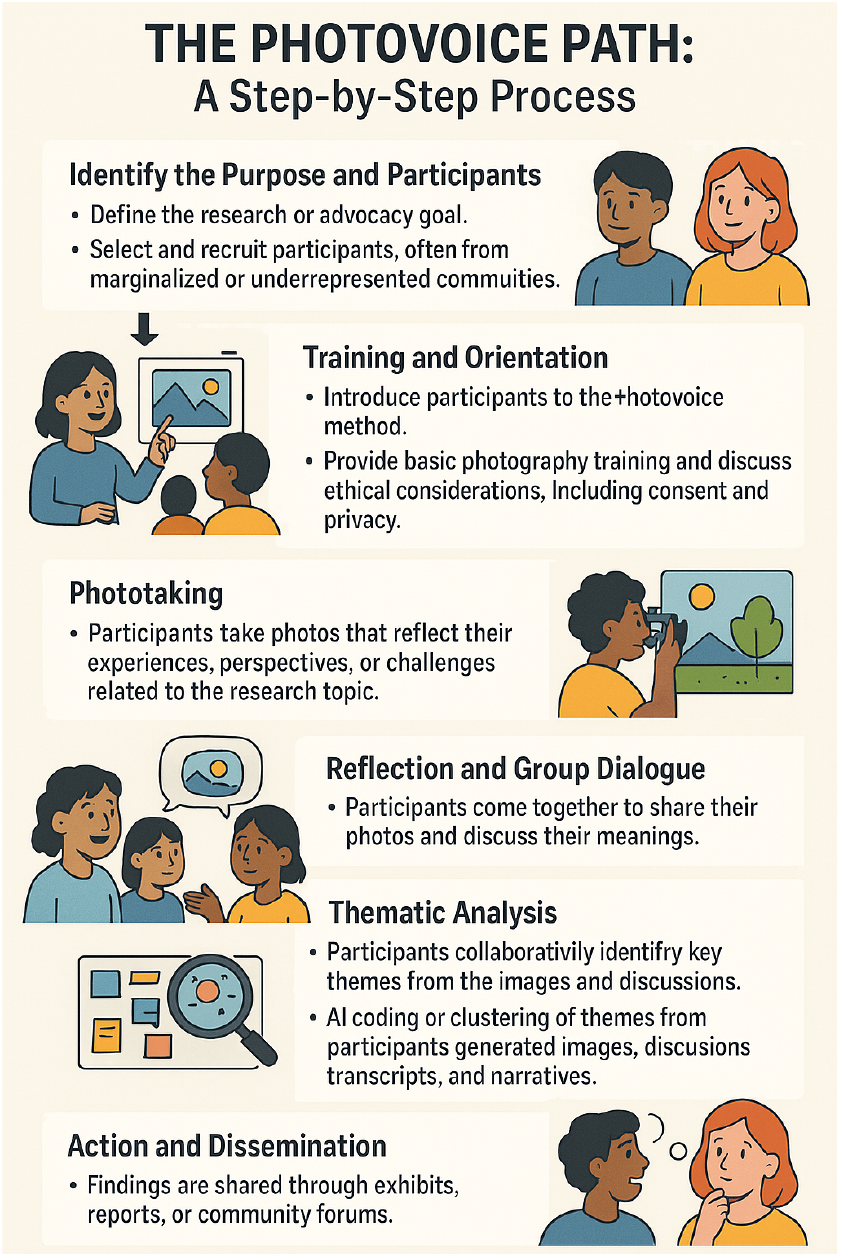
The Photovoice Path: A Step-by-Step Process (Copilot-generated)

The theoretical framework of critical theory/pedagogy has emancipatory properties and focuses on creating alternative ways for people to perceive their surroundings and circumstances. It aligns with participatory action research by centering communities as co-creators of knowledge, valuing all forms of expertise. Feminist theory further strengthens the approach by honoring subjectivity, promoting inclusive collaboration, and challenging hierarchical norms within research and policymaking (Wang et al., 1996). Photovoice is a powerful tool (Molloy, 2008), enabling the development of critical consciousness among community members through the process of photographing and discussing aspects of their daily lives (Peabody, 2013). Photovoice facilitates the creation of social spaces where people can unite to improve their collective well-being (Molloy, 2008).

### 1.2 Photovoice in medical education to address social determinants of health

Photovoice has been utilized in medical education to support critical thinking (Haffejee, 2021) in the United States and globally (Ryan et al., 2020) and to explore social justice. Photovoice provides a powerful experiential learning tool that enables students to examine social justice in a personal, reflective, and contextually-grounded manner. Social justice asserts that all individuals have inherent value and should have equal rights and opportunities. Photovoice enables medical students to move beyond social justice as a theoretical concept to an embedded aspect of their professional practice. Through taking and discussing photographs, students have critically examined how social determinants of health surround and affect communities (Andress and Purtill, 2020; Hudon et al., 2016; Loignon et al., 2014). By visualizing injustice, Photovoice helps students see the need for systemic change, supporting the development of physician-advocates who can act on what they observe.

### 1.3 The Role of AI on participatory research to advance social justice in education

Emerging evidence from studies exploring the contribution of AI techniques in participatory research and education suggest that AI has the potential to improve data analysis, understanding (Rice et al., 2025), and stakeholder engagement. AI can achieve program efficiencies including enhancing personalized medicine (Gul et al., 2024). This technological revolution redefines traditional practices, introducing new tools, methods, and ethical considerations. Photovoice is a particularly innovative integration of AI into participatory action research. Gul et al. (2024) conducted an AI-powered Photovoice analysis in sustainable consumption research, utilizing a multimodal approach that combines computer vision and natural language processing (NLP) to analyze a large dataset comprising both visual and textual elements (Gul et al., 2024). Multimodal AI integration has the potential to accelerate Photovoice analysis and enhance rigor through computational objectivity. This combination enables researchers to generate visual representations of community ideas rapidly, amplifying marginalized voices and extracting meaningful insights from unstructured data (Gul et al., 2024). For example, a high school project in Calabria, Italy (Bova, 2023), involved twenty-one students aged 17–18 in using AI-based tools, such as Midjourney, to visually depict proposals for improving local urban environments. They shared their visualizations—which addressed real-world concerns like insufficient green space—on the European platform ‘Cultural Gems.” The Calabria project depicted how AI can enhance Participatory Action Research (PAR) by making abstract ideas more generalizable and shareable.

A promising innovation is the use of conversational AI to support reflexive dialogue. Garg et al. (2024) employed a participatory AI approach to Photovoice analysis, engaging students in India as citizen scientists and documenting their perceptions and practices related to food waste(Garg et al., 2024). This method seeks to scale the application of Photovoice while maintaining a focus on participant interpretation and reflexivity. Through interactive prompts tailored to participants’ images (e.g., “What emotion do you associate with this image?”), AI can encourage deeper reflection and self-awareness.

Among a group of first-year medical students, we conducted a Photovoice study to examine the concept of social justice. We explored visual and narrative documentation of social justice based on six prompts, collaboratively analyzed the data, reflected critically on their observations and translated them into advocacy strategies relevant to their future roles in medical education.

## 2 Methodology

### 2.1 Reflexivity

We acknowledge our positionality relative to the scientific process. As the researchers conducting this participatory action research study on social justice in medical education and practice, we recognize the importance of acknowledging our positionality in shaping the research process and findings. The facilitation of the discussions, co-analysis and development of the manuscript was led by SA and SJ. All authors hold terminal degrees: MD (SA, RC) and/or EdD or PhD (SJ, DC, HRN, RC) and hold positions in academic research at the post-doctoral level or greater.

Two members of the team (SA and SJ) served as primary facilitators, guiding discussion sessions, Photovoice activities, and analytic workshops. Their dual roles as instructors and investigators inevitably shaped group dynamics, particularly in how students articulated emerging ideas. To address this, facilitators emphasized transparency, acknowledging their own perspectives and encouraging dissenting interpretations or alternative viewpoints. The team regularly engaged in reflexive debriefing after each session, documenting assumptions, shifts in understanding, and possible areas of interpretative bias. These conversations informed ongoing decisions about data collection, theme development, and integration of human- and AI-derived insights.

### 2.2 Study Context

The Photovoice sessions were conducted with nine first-year medical students who participated in a 6-week seminar as part of the medical school curriculum. Students chose the Photovoice course to explore the concept of social justice from among 12 different seminars in Narrative Medicine. The seminar provided a structured but exploratory environment for students to reflect on health care access and the broader social context of medicine.

### 2.3 Methodological orientation

Drawing from ecological and intersectionality frameworks (Slack-Smith et al., 2023), the project assessed and visually documented the perspectives of medical students on social justice.

### 2.4 Data Collection

During the first session and prior to data collection, participants received a 2-hour training session on Photovoice methodology. The training included visual literacy, communication through images, ethics in photography, and the purpose of the project. The seminar leaders and participants met for 2 to 3 hours each session to share and discuss participants’ photographs and written narratives. For the next 5 sessions, participants used their cell phones to capture photos based on a given weekly prompt spanning from the abstract to the concrete. Prompts were scaffolded to elicit both conceptual and situational reflections on justice; for example, the first prompt asked the participants to take photos of something they saw and thought others have not noticed. This prompt aimed to engage with broader ideas, sharpen their awareness, and notice their surroundings more intentionally. The investigators collected 150 images, including individual reflections and comments that contextualized each photograph.

### 2.5 Data Analysis

The data captured in the group discussions, through audio recordings and the researchers’ notes, were analyzed for themes using two analytic approaches, traditional (i.e., human-only) and computational techniques (i.e., NLP using large language models such as ChatGPT and Copilot). This hybrid analysis allowed us to identify seven initial key themes, which we then consolidated into six broad categories to achieve a more coherent and in-depth interpretation.

### 2.6 Human-only Analysis

The analysis was conducted in a 3-stage process: (1) selecting the photographs to be discussed; (2) contextualizing the photos through verbal narratives; and (3) codifying the themes, and theories that may generate meaning for a single image (Wang, 1999). The images and narratives were analyzed using the SHOWeD method (Wang et al., 1996). The participants selected meaningful photographs and explained how each image related to the weekly prompt, while the seminar leaders/investigators captured emergent insights during group dialogue, documenting both individual and collective reflections (**Table 1**). All sessions were audio-recorded and transcribed using Sonix, an AI-enabled transcription program. Transcripts were manually reviewed by investigators for accuracy before analysis.

**Table 1.** Selected insights (total 88)

| Selected insights |  |
| --- | --- |
| metaphoric | Realities |
| equal opportunities | build trust to provide care |
| a metaphor of what social justice should be | what are the implications of having less resources? |
| the bridge helping a lot of people | Inaccessible |
| everyone going to the same rate | social justice - not only an understanding of who you serve but why are we serving - those views |

At the end of the six-week seminar, the investigators displayed the photographs and insights generated during the group discussions and photography analysis on a table (**Figure 2**). The interactive display of the photographs served as a visual tool that helped the students collaboratively cluster and interpret the insight. This process produced seven themes: Unspoken Luxury, Distribution, Irony, contradictions, hypocrisy, Power, Privilege, Environmental justice, and Community Resilience.

**Figure 2.**
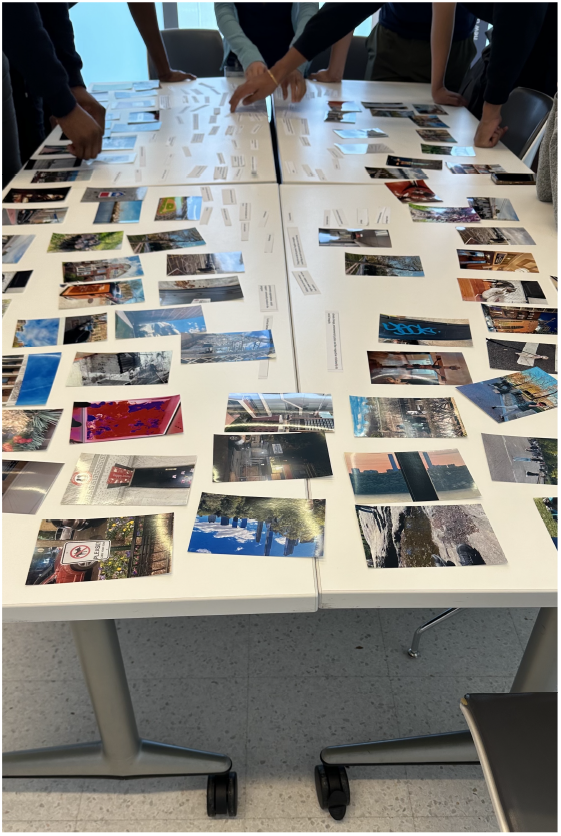
Photography and Theme Analysis

The goal was to allow the participants to work together to curate the photos based on the displayed insights and create themes and categories. The session ended with individual reflections on the overall project.

Following the final seminar session, the research team conducted a comprehensive, human-only analysis of the photographs, transcripts, written reflections, and field notes. Three investigators (SA, SJ, DC) independently performed initial inductive coding, identifying meaningful units of text—such as phrases and narrative segments—that conveyed students’ perceptions. As the analysis progressed, these codes were grouped into broader conceptual categories through thematic analysis. Using the constant comparative method (Glaser and Strauss, 1967), the team refined patterns and identified common themes grounded in the data until saturation was reached. These findings were then compared to the seven major categories that had emerged from the final group discussion and ultimately refined into six major categories including an additional theme not specifically identified by the participants (i.e. points of view), but widely discussed in group sessions. Throughout the process, particular attention was given to students’ reflections that described personal experiences, highlighted observations of disparities, and explored potential change within both medical institutions and the surrounding communities.

Participants’ words, which most faithfully conveyed the essence of their stories, were used to title the photo groupings and associated themes. To ensure rigor, the team applied multiple strategies. Credibility was supported through member checking, reflective memos, sustained participant engagement, and participants’ review of emerging themes. Dependability was strengthened by a detailed audit trail documenting coding iterations and analytic decisions. Confirmability was upheld through triangulation of data sources, photographs, narratives, transcripts, and field notes, which captured individual and group insights., as well as transparency in analytic choices and preservation of participants’ language in theme labels. Insights emerging from photo narratives were also compared with transcript-based thematic codes to confirm patterns or identify divergent perspectives. Member checking in the final session allowed participants to validate or challenge thematic insights, enhancing confirmability.

### 2.7 GenAI-based Analysis

Once participants identified key categories and selected photographs, the investigators utilized pre-selected AI tools to generate themes and visual representations based on participants’ narratives, photographs, and group discussions. Narratives and discussion transcripts were analyzed using Copilot and ChatGPT to assist in thematic coding and interpretation. For each GenAI application, we used a prompt, “analyze the following qualitative data and identify the major themes of the combined narratives and transcripts,” a second prompt ask to “combine the themes into 6 main categories to match the number of themes resulting from the human-only analysis.”

The outputs generated by these AI tools were independently reviewed by the researchers and compared with human-only analysis to identify points of convergence and divergence such as contradictions, inequity, and privilege. Researchers examined the GenAI analysis for coherence, accuracy, and potential bias. These insights were integrated into the final thematic structure, as summarized in **Table 2**. Throughout the process, AI-assisted analysis functioned only as a secondary tool, with human interpretation remaining central. This multi-method approach enhanced analytic triangulation and strengthened transparency in how the final themes emerged.

**Table 2.**
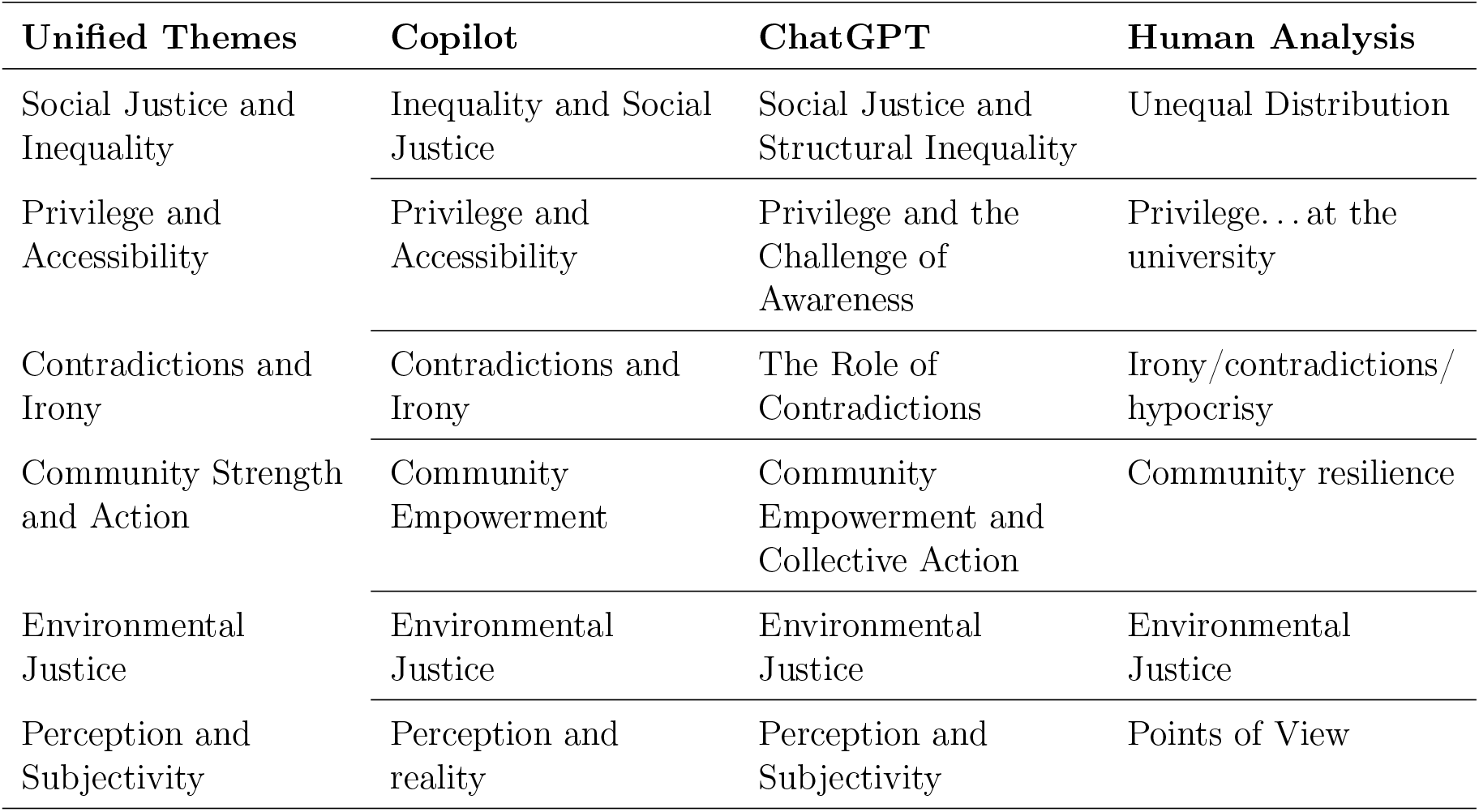
Unified Themes: Comparing Human and AI Analyses of Participant Reflections.

| Unified Themes | Copilot | ChatGPT | Human Analysis |
| --- | --- | --- | --- |
| Social Justice and Inequality | Inequality and Social Justice | Social Justice and Structural Inequality | Unequal Distribution |
| Privilege and Accessibility | Privilege and Accessibility | Privilege and the Challenge of Awareness | Privilege...at the university |
| Contradictions and Irony | Contradictions and Irony | The Role of Contradictions | Irony/contradictions/hypocrisy |
| Community Strength and Action | Community Empowerment | Community Empowerment and Collective Action | Community resilience |
| Environmental Justice | Environmental Justice | Environmental Justice | Environmental Justice |
| Perception and Subjectivity | Perception and reality | Perception and Subjectivity | Points of View |

## 3 Results

Six themes emerged from the students’ description of social justice: Social Justice and Inequality (Unspoken luxury, Distribution), Contradictions and the Costs of Justice, Community and Collective Action, Environment and Environmental Justice, Perception, Subjectivity, and Perspective. From the cross-analysis of Copilot, ChatGPT, and traditional methods, six unified thematic themes were synthesized (**Table 2**). The photographs and participant quotes provide context for each theme.

### 3.1 Theme 1: Social Justice and Inequality of Distribution

This theme brings together concerns about the unequal distribution of resources, opportunities, and rights. It includes concepts such as structural determinants of health, access to education and essential services, and the effects of social privilege. Across the analyzed platforms, narratives emerged that directly link inequality to systemic injustice, challenge the idea of meritocracy, and shed light on historical exclusions. One of the participants in this research shared a photo (see **Figure 3**) and describes the experience as follows:

**Figure 3.**
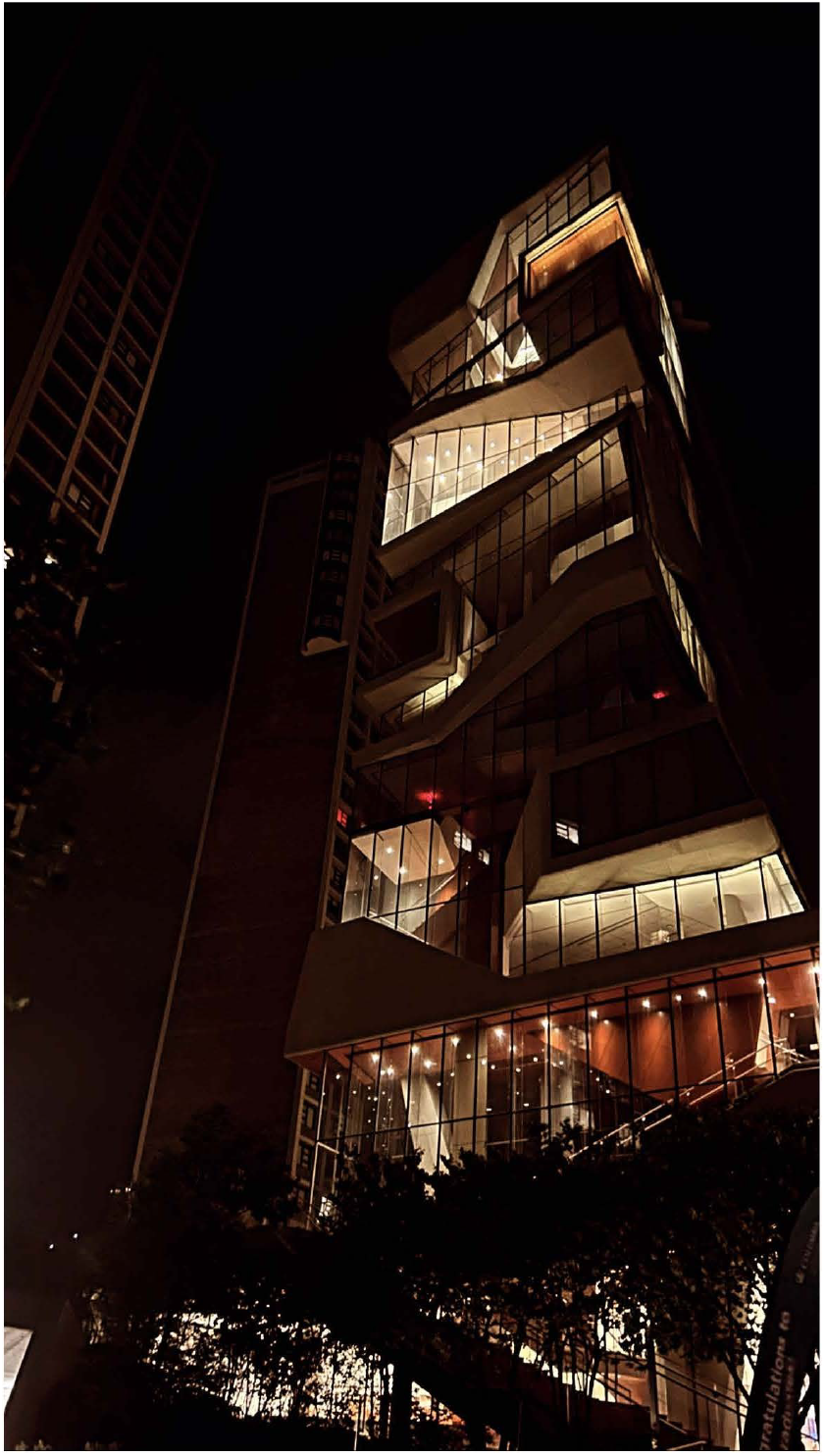
Social Justice and Inequality (Distribution - Participant’s image in the Social Justice Seminar) The image showcases the medical school building’s lights shining brightly (a recently built high rise for medical education in the middle of a working-class neighborhood and 3-5 story apartment buildings), even though it remains unoccupied during this late hour. The symbolism captured in this photo also resonates with our ongoing conversations centered around this medical education building. It stands out as a contradiction to the surrounding buildings, which reflect the disconnect between the university and the neighborhood. We have also explored various topics related to the medical building, including how this building can sometimes give the impression of looking down upon the rest of the community, as well as the power dynamics between the university and the neighborhood. In a way this reminds me of the unequal distribution of resources, which is a central concern within social justice.

### 3.2 Theme 2: Privilege

The photographs and narratives showed the participants’ perceptions of privilege, which is often invisible to those who possess it but profoundly consequential for those who do not. The group spent time discussing unspoken luxury and noted that these themes were visible in several photographs depicting privilege. **Figure 4** represents one participant’s photos and narratives:

**Figure 4.**
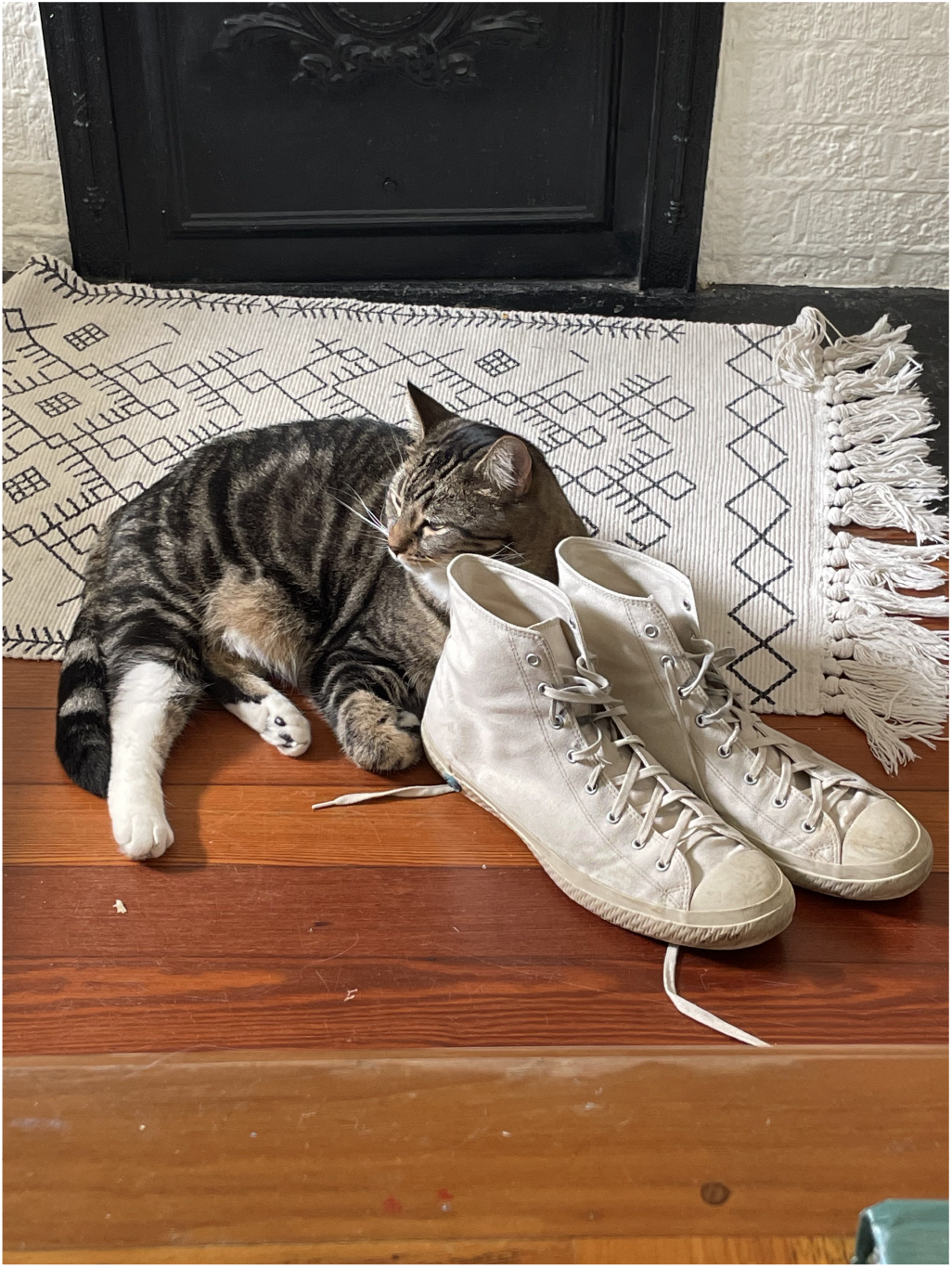
Privilege - Participant’s narrative and image in the Social Justice Seminar. I think of pets often when it comes to social justice with an almost **intangible element of privilege**. The pets in my life truly give me so much joy and comfort, but in many ways, they cost a significant amount and are not feasibly a component of many peoples’ lives, even if they could benefit from company at home or a form of healthy distraction from stressors.

### 3.3 Theme 3: Contradictions and Irony

This category encompasses internal tensions, paradoxes, and personal sacrifices involved in the pursuit of social justice. It reveals contradictions between values and practices, as well as reflections on the emotional and social toll of acting in alignment with ethical ideals. It highlights the complexity of political action within unequal contexts. The theme of contradictions captures the tension between ideals and reality, where societal promises of equity or inclusion clash with actual practices. Irony and hypocrisy emerge as ways to describe these discrepancies, revealing how systems that claim to be fair may still perpetuate injustice. The photo (**Figure 5**) and the corresponding narrative below highlight inconsistencies in behavior, policy, and institutional rhetoric.

**Figure 5.**
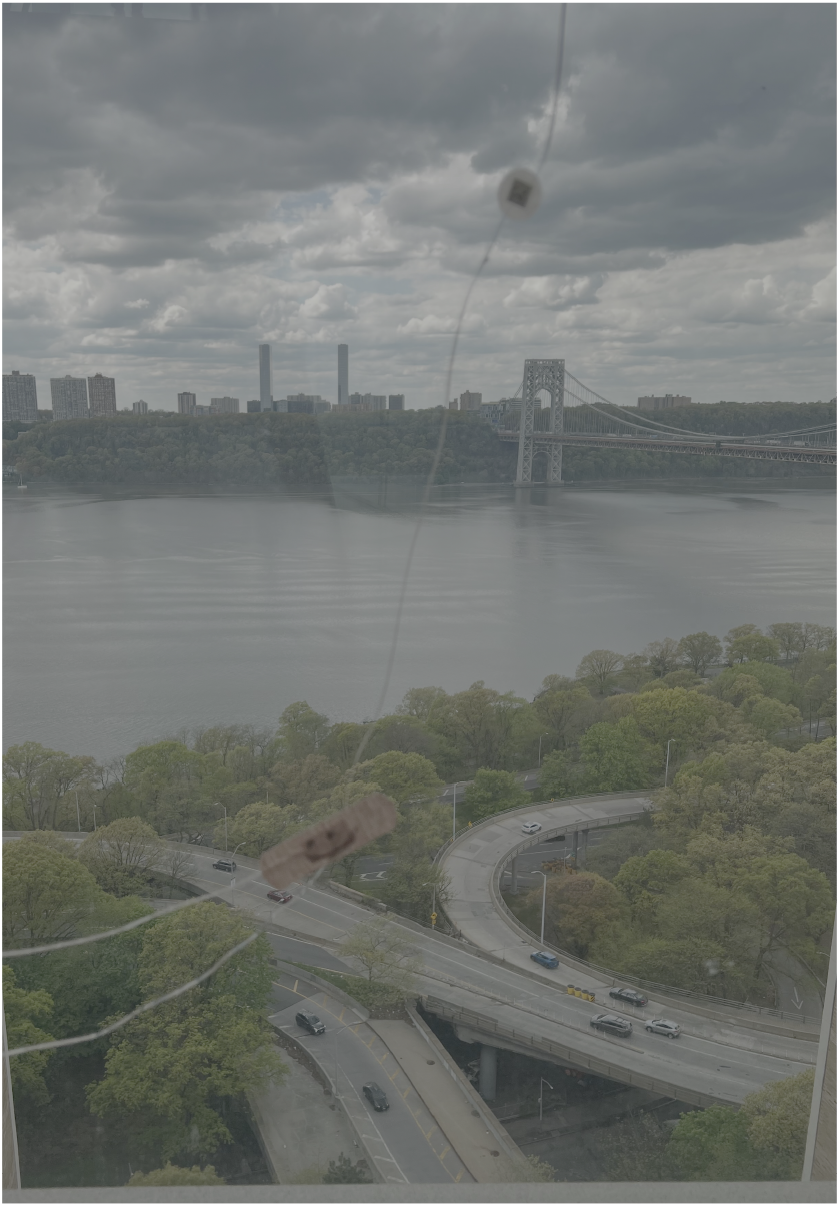
Contradictions and Irony. This picture is a crack in the glass in the medical school towers which someone put a band aid over. I have learned that it is often easy to sit complicit to injustice or not recognize how our actions and inactions contribute to the harm or marginalization of others. However, I feel like as I seek to continue my political and human education, I find it harder and harder to act like everything is normal when everyday people are being incarcerated, dehumanized, killed. This coming of awareness is a privilege and a constant pursuit; however, I have learned that attention to who is excluded, who is harmed, who is in control is imperative and extremely disconcerting at times… Every day we all apply our own little band aids to try to cover the major cracks that exist within our society. Yet those cracks represent more than just a disruption of the beautiful river view.

### 3.4 Theme 4: Community and Collective Action

This axis emphasizes the role of community as a space for resistance, mutual support, and collective building. It explores topics such as empowerment, resilience, and active participation in social transformation processes. The photo (**Figure 6**)^1^ and narrative below illustrate how collective action can arise from shared experiences of oppression, generating solidarity and agency.

**Figure 6.**
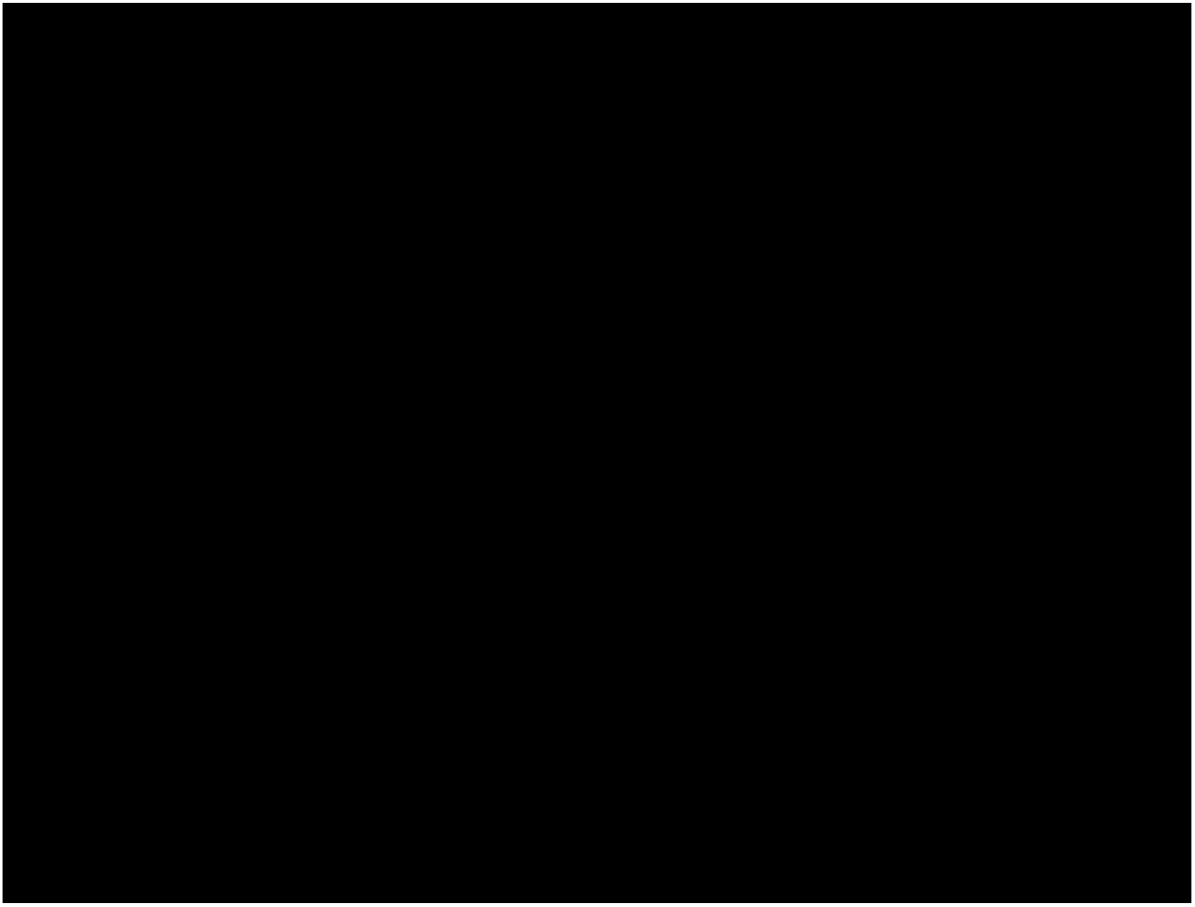
Community and Collective Action. The act of cleaning the streets by a local community organization demonstrates community empowerment and collective action. When individuals come together to clean up their neighborhoods, they are taking an active role in shaping their environment. This sense of empowerment and collective action is also essential to increased civic engagement and social cohesion, ultimately important components of promoting social justice.

### 3.5 Theme 5: Environmental Justice

This theme examines the intricate relationship between human systems and the natural world, highlighting themes of resistance, adaptation, environmental justice, and human influence. Through symbolic imagery—a leaning tree reaching for sunlight, a purple tree marked with “Salvar el planeta,” a raccoon carrying a Popeyes bag, and a heavily pruned tree—it critiques the impact of capitalism, consumerism, and aesthetic norms on nature. At the same time, it emphasizes nature’s enduring ability to adapt, persist, and subtly resist these systemic forces. This represented theme by one the participants’ narrative and photo (**Figure 7**) recognizes environmental justice as an extension of social justice as described by one of the participants:

**Figure 7.**
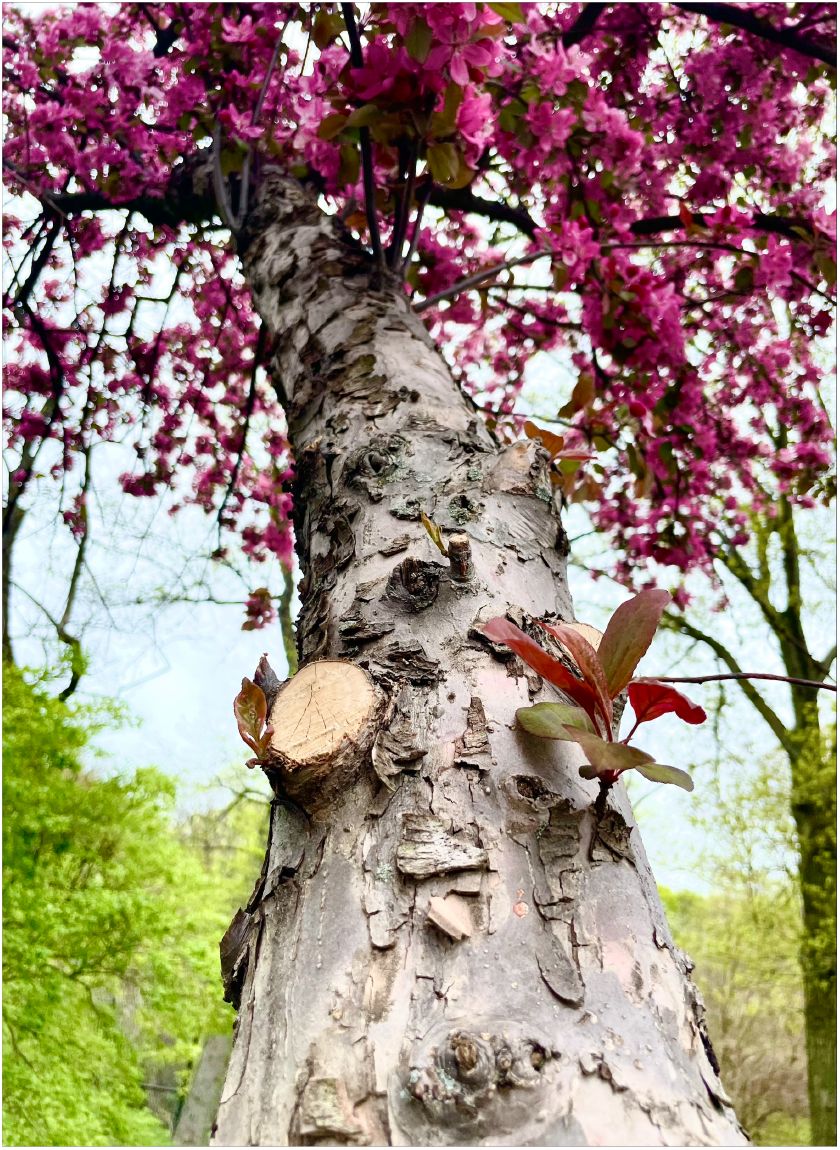
Environmental Justice. Environmental justice, climate change, and their respective relationships to social inequity is a reality we need to begin looking at more and more closely in the coming decades. I noticed that many of the branches (in the photo) had been cut and the tree manicured in many ways to make it appear more aesthetically pleasing to our eyes. It made me think of the natural things that we see that are manicured and curated for our own purposes, inadvertently harming the actual biology’s natural state.

### 3.6 Theme 6: Perception, Subjectivity, and Perspective

This theme examines how individual perspectives influence the understanding of justice, inequality, and lived experiences. It recognizes that people interpret the same reality in different ways, influenced by their cultural, emotional, and social positions. These subjective viewpoints generate multiple truths, underscoring the importance of dialogue and empathy in fostering a shared understanding. The photo (**Figure 8**) and narrative below illustrate this theme:

**Figure 8.**
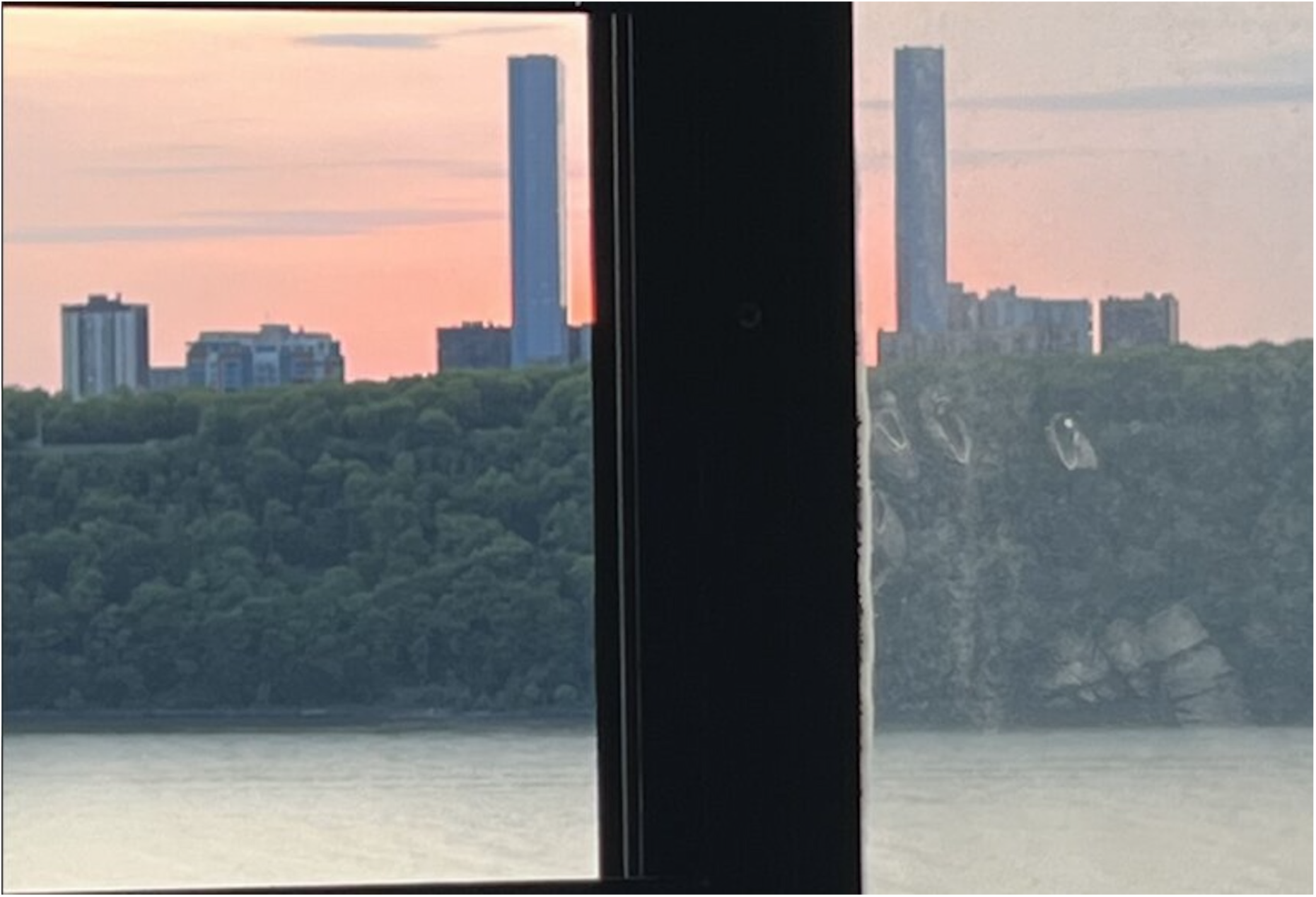
Perception, Subjectivity, and Perspective. The photo of two identical buildings don’t look the same based on the side from which they are viewed. One’s viewpoint does not always equal another’s. Reality can be distorted if the medium through which one views the same thing is clouded.

## 4 GenAI-based Analysis

Additionally, the research team used ChatGPT, Copilot, ImagineArt, Midjourney, deepai.org Design.ai, and Imagine.art (ultrarealism) to generate visual representations based on 2 participants’ narratives. Here we discussed only 2 out 150 participants’ photos and narratives that illustrate the comparative analysis that we conducted. We compared the AI-generated visuals with the original photographs taken by participants to explore differences in representation and emotional depth paying particular attention to the potential bias in the AI generated images. **Figure 9** shows a comparison between the photograph captured by the participant and the image created by AI.

**Figure 9.**
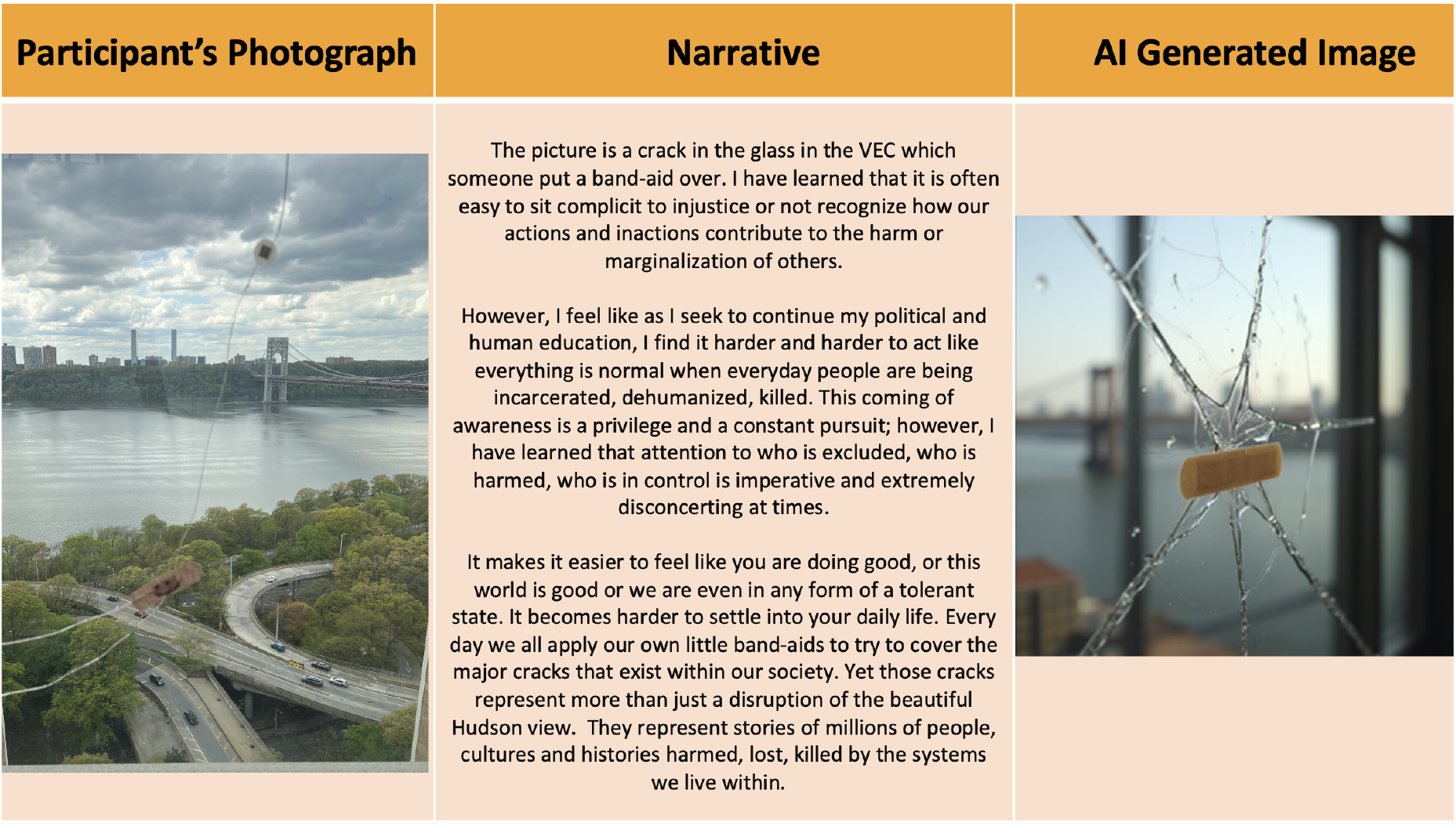
Comparison of Participant’s Photo with an AI-Generated Image from a Photo Narrative

The following **Figure 10**^2^ shows a comparison of participants’ photos and AI-generated images with different AI-powered design tools.

**Figure 10.**
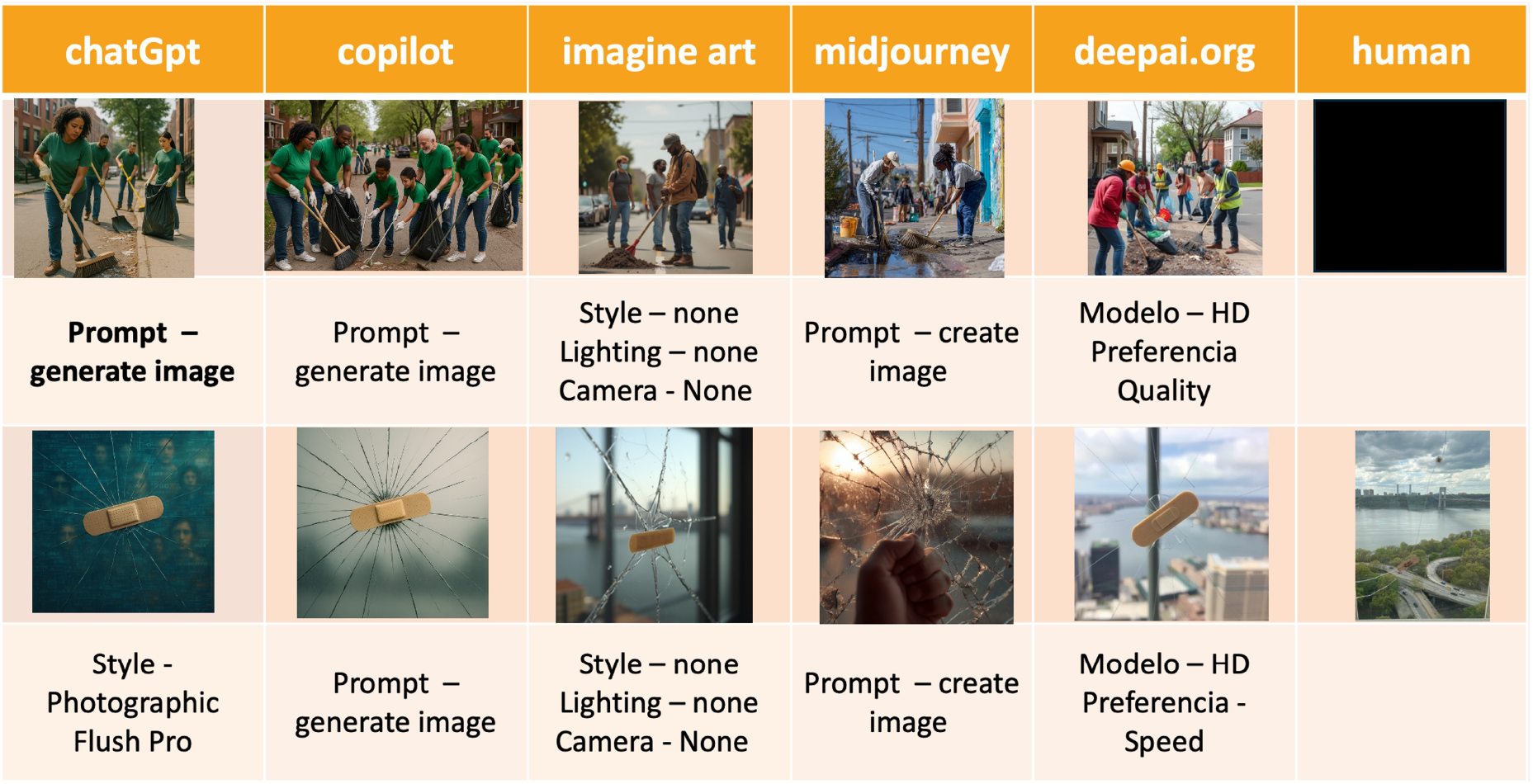
Comparison of Participants’ Photos with AI-Generated Images from Participants’ Narratives

## 5 Conclusions

This participatory Photovoice study, supported by AI-powered analysis of text and images, examined medical students’ perceptions, practices, and understandings of social justice. Rooted in a critical and transformative research tradition, the study approaches social justice through an intersectional lens, emphasizing not only equitable resource distribution but also cultural recognition, political representation, and the inclusion of marginalized voices (Fraser, 2009; Young and Nussbaum, 2011).

Thematic analysis was contextualized, acknowledging the role of persistent inequalities. Participant-generated photographs and narratives offered emotionally rich insights into justice work, revealing that efforts toward equity often entail personal sacrifice—of comfort, safety, or opportunity—especially for those actively resisting oppressive systems. Central themes such as irony, the costs of advocacy, and the connection between inequality and community-based resistance emerged as powerful framing devices.

Privilege is identified as a structural advantage linked to race, class, gender, and citizenship status (McIntosh, 1989; Kimmel, 2014). Recognizing it requires unpacking normalized disparities and making visible the systemic barriers that often go unacknowledged. Empowerment frameworks emphasize how lived experiences fuel social transformation (Freire, 1970; Zimmerman, 2000), framing community resilience as an act of resistance and collective memory, not merely passive endurance.

Finally, this article contributes to the growing discourse on the role of social justice in medical education and the integration of AI in research and pedagogy. It highlights how medical educators can utilize the Photovoice methodology to help students capture and share their lived experiences of social justice, fostering dialogue and uncovering sources of resilience.

The findings reveal a meaningful complementarity between traditional and AI-assisted approaches in qualitative visual research. The genAI-supported analysis yielded themes closely aligned with the participants’ analysis and core issues of social justice. Participant-generated Photovoice images convey emotionally rich, context-specific narratives rooted in lived experience, making them powerful tools for advocacy and authentic storytelling. In contrast, AI-generated visuals offer efficiency, adaptability, and the ability to identify subtle, latent patterns that might elude human analysis, thereby potentially enhancing generalizability and reducing researcher bias. However, they often lack the emotional depth and contextual nuance that human-created content possesses.

When used thoughtfully, these methods can enrich one another: traditional approaches anchor research in authenticity and emotional resonance, while AI brings scalability, consistency, and sophisticated pattern recognition. Their integration, particularly within participatory and reflexive frameworks, enables more profound and more comprehensive insights—especially in complex or data-intensive environments.

AI is often framed as neutral and objective. However, its inputs, design, and outputs are deeply entangled with the cultural values, social hierarchies, and historical inequities of the world it reflects. Rather than erasing prejudice, AI frequently reproduces and intensifies it, under the guise of rationality and efficiency. Scholars such as Bender et al. (2021) highlight how these data and algorithms reflect social hierarchies rather than correcting for them. Without interpretation of results, AI systems risk becoming mirrors of existing social biases and systemic inequalities (Bender et al., 2021).

Bias within AI systems stems from many interconnected sources such as training data shaped by structural inequality, developer decisions influenced by dominant norms, and a lack of sociocultural context in design (Benjamin, 2019). In his book the author describes how emerging technologies can amplify bias through design. AI models reflect the values, assumptions, and blind spots of those who build them. If design teams lack diversity or critical awareness, systems may reinforce dominant worldviews while marginalizing others (Noble, 2018). Take, for instance, the AI-generated images intended to illustrate community action: the algorithm defaulted to depicting people of color. On the surface, such representation may appear inclusive.

Ultimately, AI cannot fully comprehend the context, culture, and emotions of humans. It identifies patterns but not meaning. In research involving identity, inequality, or justice—such as Photovoice—this is a severe limitation (Boyd and Crawford, 2012; Floridi and Cowls, 2019). Bias in AI is not just a technical issue; it is a social one. It requires users to remain aware of potential bias, reflect on and analyze responses, and engineer prompts thoughtfully to avoid reinforcing existing prejudices.

### 5.1 Students’ reflections

The Photovoice process also sparked critical reflection and agency among participants. Many expressed a heightened awareness of their beliefs in social justice and a determination to change. The reflections show a personal and collective journey toward an understanding of social justice. They emphasize how awareness evolves, not through theory alone, but through lived experience, dialogue, and intentional reflection. Participants came to recognize the often-overlooked manifestations of injustice, and the prevalent nature of inequity embedded in everyday life. At the same time, the reflections highlight the power of community resilience and grassroots action as vital forces for change and how paying attention to subtle details and shared struggles fosters a richer, more grounded commitment to equity and justice, transforming limited understanding into conscious, sustained engagement, as one student reflected:

> *Prior to participating in this class, I didn’t have a strong grasp on what social justice truly was. I had sat in many institutional lectures where the concept of social justice was the main topic, but I always felt like I was listening to very generalized statements that had no meaning. Every session left me with more to be desired. This class provided me the opportunity to converse with classmates and listen to experiences, opinions, stories, and overall have a better understanding of what social justice means to me and to my peers. From the first assignment of finding things that would be unnoticed by most people, I didn’t see the connection to social justice initially. Now, I understand how this was a metaphor to many social justice issues; most people don’t pay attention. Throughout the class, I began listening to the many insights and stories my classmates would bring every week. Whether it was the many instances of hierarchy over the community through the medical school high rises, the buildings of billionaire’s row towering over central park, or the failed attempts at having social justice conversations at fancy dinners. I learned of how easily social injustice was seen, and how rare social justice was found. Overall, I’m leaving this class with a much more concrete definition of what social justice means to me, and a better awareness of the social injustices that surround me*.
>
> *Our work in this class has challenged me to think about the cost of social justice. This crucifix a symbol of Christianity, also challenges me to think about the cost of social justice The Christian worldview is that the world is good but imperfect and unjust, and the crucifix symbolizes that justice is available to everyone but comes at a great cost. I think back to the pictures of the GWB [bridge] traffic, the band-aid on the window crack, and the basketball court, and I think of the costs of solutions to big problems. What sacrifices are we willing to make to achieve justice? At this point in our education, we’re exploring and deciding what we want our careers to do for the world. I’ve been reflecting a lot on what costs I’m willing to pay and what sacrifices I’m willing to make*.

### 5.2 Considerations

One innovative approach that supports reflection and social change is the use of Photovoice as a research methodology. Photovoice allows participants to document and share their lived experiences through photography, fostering a deeper understanding of complex social issues. This method encourages individuals to reframe their self-perception and that of their communities, fostering dialogue and collaboration among researchers, participants, and policymakers.

By amplifying marginalized voices, Photovoice promotes transformation—not only in how social issues are perceived but also in how they are addressed. It bridges personal experience with collective action, generating insights that can inform practice and policy to achieve greater social justice.

By applying computer vision techniques, the content and composition of participant photos can be analyzed to identify recurring visual elements—such as symbols, settings, or objects—that are emotionally or politically charged (Radford et al., 2021). These visual cues can then be correlated with participants’ narratives to build a more emotionally rich and contextually grounded understanding.

Future research should focus on developing frameworks that support the integration of AI with qualitative approaches. Researchers can develop hybrid models that enrich participatory inquiry by combining the interpretive depth of traditional methods with AI’s pattern recognition capabilities. This synergy opens new possibilities for engaging learners, promoting critical reflection, and expanding research methodologies in socially conscious and inclusive ways.

## Data Availability

All data produced in the present study are available upon reasonable request to the authors.

## Ethical Approval

Ethical approval was obtained from Columbia University Irving Medical Center Research Ethics Committee (RASCAL #AAAV7662).

## Informed Consent

Informed consent was obtained from all participants involved in the study, ensuring respect for confidentiality and voluntariness principles.

## Footnotes

1 MedRxiv does not post articles that include images that could identify an individual, even when faces are obscured, blurred, covered, or pixelated. To comply with medRxiv policy, we removed Figure 6 from the pre-print version of this manuscript. The photograph depicts a group of volunteers, some wearing light blue t-shirts, gathered on a city sidewalk for what appears to be a local cleanup event.

2 MedRxiv does not post articles that include images that could identify an individual, even when faces are obscured, blurred, covered, or pixelated. To comply with medRxiv policy, the pre-print version of this manuscript does not display the image embedded in the first row of the last column. The participant photo depicts a group of volunteers, some wearing light blue t-shirts, gathered on a city sidewalk for what appears to be a local cleanup event.

